# Prospective evaluation of a self-report–guided strategy for targeted coronary artery calcium imaging

**DOI:** 10.64898/2026.08.17.26360575

**Authors:** Eva Hagberg, Elias Björnson, Martin Adiels, Bledar Daka, Louise H. Fornander, Josefin Kjelldahl, David Molnar, Carlo Pirazzi, Ulf Strömberg, Gustav Kjellsson, Carl Bonander, Mikael Svensson, Anders Gummesson, Göran Bergström

**Affiliations:** Department of Molecular and Clinical Medicine, Institute of Medicine, Sahlgrenska Academy, University of Gothenburg, Gothenburg, Sweden; Department of Clinical Physiology, Sahlgrenska University Hospital, Gothenburg, Sweden; School of Public Health and Community Medicine, Institute of Medicine, Sahlgrenska Academy, University of Gothenburg, Gothenburg, Sweden; Primary Health Care Center, School of Public Health and Community Medicine, Institute of Medicine, Sahlgrenska Academy, University of Gothenburg, Gothenburg, Sweden; Department of Radiology, Sahlgrenska University Hospital, Gothenburg, Sweden; Department of Cardiology, Sahlgrenska University Hospital, Gothenburg, Sweden; Centre for Societal Risk Research, Karlstad University, Karlstad, Sweden; Centre for Health Governance, Department of Economics, School of Public Health and Community Medicine, University of Gothenburg, Sweden; Department of Clinical Genetics and Genomics, Sahlgrenska University Hospital, Gothenburg, Sweden

## Abstract

**Background:** Coronary artery calcium (CAC) imaging directly assesses subclinical calcified coronary atherosclerosis, but population-wide imaging is not recommended. Simple pre-screening may help identify individuals most likely to benefit from CAC imaging. We previously developed a self-report-based model to estimate the probability of CAC ≥100. This study prospectively evaluated a strategy based on this model to select individuals for CAC imaging. We assessed agreement between model-predicted probability and observed prevalence of CAC ≥100 among participants undergoing computed tomography (CT) imaging and examined patterns of preventive lipid-lowering therapy.

**Methods:** The PRedict and Identify cOronary atherosclerosis–Now (PRIO-Now) study applied a prospective, two-step, population-based screening approach. Individuals aged 59– 60 years were invited to complete a self-report questionnaire. Eligible respondents without previous ischemic heart disease whose model-predicted probability of CAC ≥100 exceeded the predefined threshold were invited to clinical assessment and non-contrast coronary CT imaging. The primary analysis assessed agreement between model-predicted probabilities and the observed prevalence of CAC ≥100 among CT completers.

**Results:** Of 8,000 invited individuals, 2,588 (32%) completed the questionnaire. Of 2,375 eligible respondents, 814 were classified as high risk and 563 underwent CT imaging. Among CT completers, the mean predicted probability of CAC ≥100 was 28.3% (95% CI 27.2–29.3), compared with an observed prevalence of 28.4% (95% CI 24.8–32.4), corresponding to an expected/observed ratio of 0.99 and a Brier score of 0.19. Among participants with CAC ≥100, 64% were not receiving lipid-lowering therapy and 11% had LDL-C ≤1.8 mmol/L.

**Conclusions:** A self-report-guided strategy enabled targeted CAC imaging in a model-selected cohort. Among participants completing CT imaging, the observed prevalence of CAC ≥100 was comparable with the mean model-predicted probability. These findings suggest that self-report data may support pre-selection for CAC imaging and help identify opportunities for preventive treatment among individuals with elevated CAC.

**What is already known on this topic:** Coronary artery calcium (CAC) is a direct measure of subclinical coronary atherosclerosis and refines cardiovascular risk assessment beyond traditional risk factors. However, population-wide CAC screening is not recommended, and current risk-based selection approaches may miss individuals with prognostically relevant disease.

**What this study adds:** This prospective evaluation assessed a previously developed self-report-based model for identifying individuals with moderate to severe coronary atherosclerosis (CAC ≥100). In the model-selected high-risk group, the predicted probability of CAC ≥100 was similar to the observed prevalence among participants completing CT imaging. The study also showed that many individuals with CAC ≥100 were not receiving lipid-lowering therapy.

**How this study might affect research, practice or policy:** Self-report-based pre-screening may support more selective use of CAC imaging in future prevention strategies. By identifying individuals with substantial subclinical coronary atherosclerosis, this approach may help detect treatment opportunities and guide more targeted preventive intervention.

## Introduction

Coronary heart disease (CHD) remains a leading cause of morbidity and mortality worldwide despite advances in preventive therapy.^1^ Assessment of coronary artery calcium (CAC) in Agatston units by non-contrast computed tomography (CT) improves cardiovascular risk stratification beyond traditional risk factors and helps identify individuals who may benefit from preventive therapy.^2-6^ Despite its clinical value, CAC imaging is limited by cost and accessibility; radiation exposure with modern low-dose CT is minimal.^7^ Current prevention guidelines primarily rely on traditional risk scores to guide selection of individuals who may benefit most from CAC imaging, typically those at intermediate risk or close to treatment decision thresholds.^10,11^ To improve the efficiency of CAC screening, self-report–based pre-screening approaches have been proposed to reduce unnecessary imaging while identifying individuals with elevated CAC and CHD risk.^8-11^

In previous work, we developed a self-report model within the Swedish CardioPulmonary BioImage Study (SCAPIS) to identify individuals with CAC ≥100.^9^ In cross-sectional analyses, the model showed good discrimination for CAC ≥100 (AUC 0.81), comparable to more comprehensive models and higher than the Pooled Cohort Equations (AUC 0.75).

To prospectively evaluate this self-report-guided screening strategy in a population-based setting, we designed the PRedict and Identify cOronary atherosclerosis-Now (PRIO-Now) study. The primary aim was to assess agreement between model-predicted probability and observed prevalence of CAC ≥100 among participants completing CT imaging. A secondary aim was to examine use of lipid-lowering therapy among individuals with elevated CAC.

## Methods

### Study Design

The PRIO-Now study was a prospective, observational cohort study conducted in two steps, an initial web-based questionnaire (step 1) followed by clinical assessment and CAC imaging (step 2). Individuals who completed the web-based questionnaire and were classified as high risk by the model were invited to clinical assessment and imaging. All visits were scheduled within six months of the web-based questionnaire.

The prediction model used in this study was developed in the Swedish CardioPulmonary BioImage Study (SCAPIS; n=30,154), a nationwide population-based cohort with standardized cardiac CT, clinical examinations, and questionnaires.^9^ In this manuscript, it is referred to as the PRIO-Now model and was applied without modification.

The study was approved by the Swedish Ethical Review Authority (Dnr 2023-03227-01), and all participants provided informed consent. The study adhered to the Declaration of Helsinki.

### Data Availability

Due to the sensitive nature of the data, access is restricted. However, access to data, analytical methods, and study materials can be arranged upon reasonable request to the corresponding author, in line with Swedish legislation.

### Recruitment of participants

Individuals aged 59–60 years were randomly selected from the Swedish population register within the greater Gothenburg catchment area, with equal numbers of women and men invited. The register identifies all registered residents through unique personal identity numbers, enabling direct population-based sampling. The narrow age range was chosen to reduce age-related variation in CAC prevalence and cardiovascular risk and to evaluate the screening strategy in a clinically relevant age group for preventive intervention.

A total of 8,000 invitation letters were sent by postal mail, inviting recipients to provide informed consent online and complete a web-based questionnaire. Exclusion criteria, based on self-reported information, included history of myocardial infarction (MI), coronary artery bypass grafting (CABG), percutaneous coronary intervention (PCI), and angina. Participants with blank or near-blank questionnaires were excluded, as were participants with incomplete contact information.

### Web-based questionnaire

The questionnaire consisted of 14 self-assessed items covering demographic and anthropometric factors (sex, age, weight at age 20, current weight, height, waist and hip circumference), smoking history (current smoking status and pack-years), medical history (use of lipid-lowering or antihypertensive medications, diagnosed diabetes, diagnosed hypertension), and family history of MI (family history of myocardial infarction before age 60 in a first-degree relative). The PRIO-Now model uses the self-assessed questionnaire variables to estimate the probability of having CAC ≥100.^9^ Key differences between the self-report model and traditional risk scores are presented in Supplemental Table S1.

A predefined risk threshold of 15.3% defined high risk and was derived from the SCAPIS cohort used to develop the model, corresponding to the 30% of individuals at highest-risk, aged 59–60 years. This threshold captures approximately 60% of individuals with CAC ≥100.^9^ (see Supplemental Methods for details). It was selected to balance the number of individuals referred for CT imaging with the ability to identify a substantial proportion of individuals with CAC ≥100. Participants with a predicted risk ≥15.3% were invited to clinical assessment and CT imaging.

### Clinical assessment and CAC imaging

Participants classified as high risk were invited to attend two scheduled visits.

At the first visit, clinical assessments were performed including measurements of height, weight, and waist circumference, blood pressure and 12-lead electrocardiogram (see Supplemental Methods). Fasting venous blood samples were collected and analysed for low-density lipoprotein cholesterol (LDL-C), high-density lipoprotein cholesterol (HDL-C), triglycerides, and additional biomarkers. Participants also completed a detailed questionnaire. At the second visit, cardiac imaging for CAC score was performed using electrocardiogram-gated non-contrast CT at 120 kV. A specialist thoracic radiologist reviewed all scans for incidental non-coronary findings.

### Participant feedback and clinical management

Participants with predicted risk below 15.3% were classified as normal risk and received written feedback with general lifestyle advice. No further study procedures were performed.

Participants identified as high risk received a written report with CAC score, laboratory results, and clinical measurements. For participants with CAC ≥100 and CAC ≥300, recommendations were provided based on study-defined LDL-C levels of ≤1.8 mmol/L and ≤1.4 mmol/L, respectively. These levels were selected by the study group based on published guidance and expert consensus.^1,12-15^ Participants with diabetes were advised to discuss LDL-C targets with their primary care provider. Individuals with CAC ≥300 were contacted by telephone, and treatment recommendations were discussed with a cardiologist. Incidental non-coronary findings were managed and communicated according to clinical relevance.

### Outcome measures

#### Primary outcome

The primary outcome was to assess whether predicted probabilities of CAC ≥100 corresponded to the observed prevalence among participants completing CT imaging.

#### Secondary outcomes

Secondary outcomes included elevated blood pressure and glucose levels among participants attending clinical assessment. Among those undergoing CT imaging, secondary outcomes included lipid-lowering therapy use and study-defined LDL-C levels in relation to CAC category (≤1.8 mmol/L for CAC ≥100 and ≤1.4 mmol/L for CAC ≥300), as well as the frequency and type of incidental non-coronary findings.

### Statistical Analysis

All analyses were performed using R (version 4.1.3).

#### Sample size considerations

Sample size considerations were based on estimating the prevalence of CAC ≥100 among individuals aged 59–60 years classified as high risk by the PRIO-Now model. Based on prior analyses from SCAPIS, the expected prevalence in the highest-risk 30% was approximately 29%. A sample of 318 participants undergoing CT imaging was estimated to provide a two-sided 95% confidence interval with a margin of error of approximately ±5 percentage points around this prevalence estimate. To allow for uncertainty in response rates, exclusions, and non-attendance, the study aimed to recruit approximately 500 individuals for clinical assessment and CAC imaging. Assuming that approximately 30% of questionnaire respondents would meet the predefined high-risk threshold, approximately 8,000 invitation letters were distributed.

#### Participation and missing data analyses

Area-level socioeconomic differences in response rates were examined using median income data for DeSO (Demographic Statistical Areas), obtained from Statistics Sweden. Invited individuals were linked to a DeSO based on their residential information, and the areas were categorised into quartiles according to median income. Sex differences in response rates were also assessed.

Missing self-reported predictor variables used for model prediction were imputed using a K-nearest neighbour algorithm with five neighbours. No imputation was performed for clinical variables.

Baseline self-reported characteristics were compared between participants classified as high risk who attended the clinical assessment and those who did not. Among participants attending the clinical assessment, baseline clinical characteristics were also compared between those who completed CT imaging and those who did not.

#### Descriptive and comparative analyses

Continuous variables are presented as mean (SD) or median [IQR], as appropriate, and categorical variables as n (%). Group differences were assessed using chi-square tests for categorical variables, and either the Wilcoxon rank-sum test or Student’s t-test for continuous variables, as appropriate based on variable distribution. Chi-square assumptions were assessed before analysis. Overall model performance was evaluated using the Brier score, with 0 indicating perfect prediction and 1 reflecting poor predictive performance.^16^ Calibration was assessed using Spiegelhalter’s Z-test.^17^ Additional calibration metrics, including expected/observed ratio, calibration intercept, and calibration slope, were evaluated. Calibration plots comparing predicted probability with observed prevalence of CAC ≥100 were generated using tertiles and quintiles of predicted risk. Confidence intervals for observed prevalence were calculated using binomial methods. Confidence intervals for mean predicted risk were calculated from the standard error of the mean.

#### Agreement between self-reported and clinical variables

In the original SCAPIS model, waist and hip circumference were clinically assessed, whereas in PRIO-Now these variables were initially self-reported in the web-based questionnaire. To examine whether this difference in data collection influenced predicted probabilities, we performed an additional analysis among participants with available paired data. Predicted probabilities based on self-reported anthropometric variables were compared with probabilities recalculated using clinically reassessed waist and hip circumference measurements.

#### Patient and Public Involvement

Patients and the public were not involved in the design, conduct, reporting or dissemination of this research.

## Results

### Web-based screening and model-based selection

Between September and November 2023, 8,000 individuals aged 59–60 years, of whom 50% were male, were invited to participate by postal mail. In total, 2,588 individuals (32%) responded and completed the web-based questionnaire; 46.8% of respondents were male. Response varied across area-level socioeconomic strata, ranging from approximately 15% in the lowest income quartile to 42% in the highest income quartile.

After exclusion of individuals with previous MI, CABG, PCI, angina, blank or near-blank questionnaires, or missing contact details, 2,375 eligible respondents remained. Participants were classified as having a predicted risk below or above the predefined threshold of 15.3% using the PRIO-Now model (Figure 1). Baseline characteristics stratified by predicted risk are shown in Table 1. The high-risk group had a greater burden of cardiovascular risk factors and was predominantly male (89%). Participants with a predicted risk ≥15.3% (n=814) were invited to clinical assessment and CT imaging.

**Figure 1:**
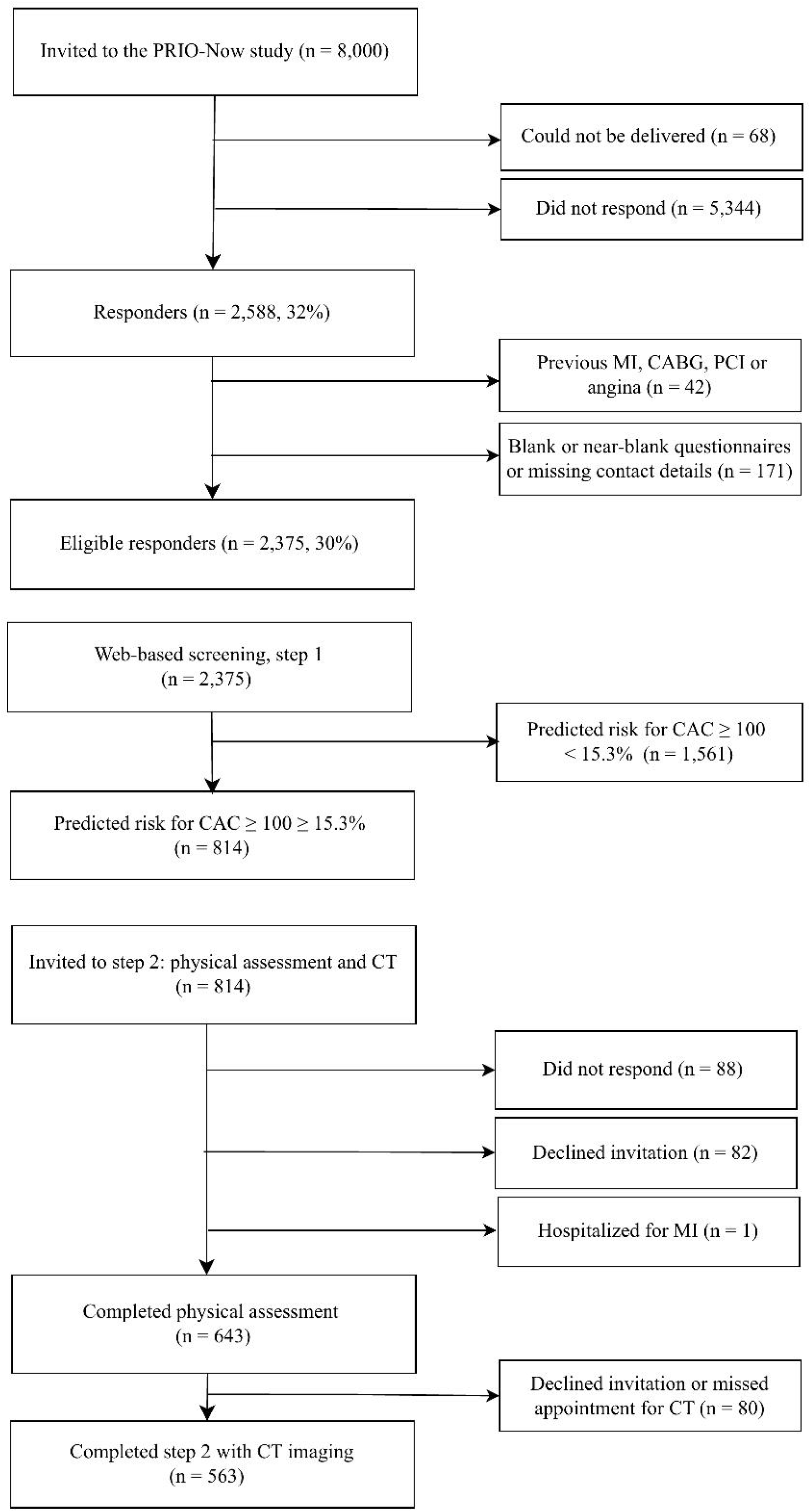
Flow diagram of the study participants. Overview of participant recruitment, web-based pre-screening (step 1), risk stratification, and clinical assessment including coronary CT imaging (step 2) in the PRIO-Now study. CABG, coronary artery bypass grafting; CAC, coronary artery calcium; CT, computed tomography; MI, myocardial infarction; PRIO-Now, PRedict and Identify cOronary atherosclerosis–Now.

**Table 1:** Baseline characteristics of 2,375 participants completing the web-based pre-screening questionnaire, stratified by predicted risk group.

| Characteristics | All<br>(n=2,375) | Predicted risk<br><15.3% (n=1,561) | Predicted risk<br>≥15.3% (n=814) | p-value |
| --- | --- | --- | --- | --- |
| Sex (n, % male) | 1,111 (46.8) | 387 (24.8) | 724 (88.9) | < 0.001 |
| Age, years | 59.4 (0.6) | 59.3 (0.6) | 59.5 (0.6) | < 0.001 |
| Body weight at age 20, kg | 66.1 (11.6) | 61.7 (9.3) | 74.1 (10.9) | < 0.001 |
| Body weight, kg | 80.2 (15.6) | 75.0 (13.1) | 89.6 (15.5) | < 0.001 |
| Height, cm | 173.2 (9.4) | 170.2 (8.5) | 178.6 (8.4) | < 0.001 |
| Waist circumference, cm | 96.5 (13.3) | 92.5 (11.7) | 104.1 (12.9) | < 0.001 |
| Hip circumference, cm | 103.5 (9.7) | 103.1 (9.6) | 104.1 (10.0) | 0.04 |
| Current smoker (%) | 102 (4.3) | 35 (2.4) | 67 (8.2) | < 0.001 |
| Cigarette pack-years | 0.0 [0.0, 5.0] | 0.00 [0.0, 3.0] | 0.0 [0.0, 13.0] | <0.001 |
| Use of lipid-lowering medication (%) | 265 (11.2) | 74 (4.7) | 191 (23.5) | < 0.001 |
| Use of antihypertensive medication (%) | 538 (22.7) | 198 (12.7) | 340 (41.8) | < 0.001 |
| Diagnosed diabetes (%) | 118 (5.0) | 26 (1.7) | 92 (11.3) | < 0.001 |
| Diagnosed hypertension (%) | 809 (34.1) | 358 (22.9) | 451 (55.4) | < 0.001 |
| Family history of myocardial infarction* (%) | 290 (12.2) | 129 (8.3) | 161 (19.8) | < 0.001 |

\* Father, mother, or biological sibling had a myocardial infarction before 60 years of age. Values are presented as n (%), mean (SD), or median [interquartile range (IQR)] as appropriate.

### Clinical attendance, missing data, and CT completion

Among the 2,375 participants with model-based risk classification, 424 (17.9%) had at least one missing self-reported predictor variable. Missingness was most common for waist and hip circumference (Supplemental Table S2). Of the 814 high-risk participants invited to clinical assessment, 643 attended the first visit and 563 subsequently completed coronary CT imaging for CAC scoring (Figure 1). Missingness in clinical laboratory variables was low (Supplemental Table S3).

Participants attending the clinical visit were more often male and less likely to be current smokers than invited high-risk participants who did not attend, although other baseline self-reported characteristics were broadly similar between groups (Supplemental Table S4). Baseline clinical characteristics were also broadly similar between participants who completed CT imaging and those who attended the clinical assessment but did not undergo CT imaging (n=80; Supplemental Table S5).

At clinical assessment, 26.3% of participants had blood pressure ≥140/90 mm Hg and 6.4% had HbA1c ≥48 mmol/mol (Supplemental Tables S6 and S7). Among the 563 participants completing CT imaging, 160 (28.4%) had CAC ≥100 and 71 (12.6%) had CAC ≥300. Baseline characteristics stratified by CAC category are presented in Table 2.

**Table 2:** Baseline characteristics of the 563 study participants completing CT scan.

| Characteristic | All<br>(n=563) | CAC score<br><100 (n=403) | CAC score<br>≥100 (n=160) | p-value |
| --- | --- | --- | --- | --- |
| **Demographics** |  |  |  |  |
| Sex (n, % male) | 500 (88.8) | 357 (88.6) | 143 (89.4) | 0.905 |
| Age, years | 59.5 (0.6) | 59.5 (0.6) | 59.5 (0.6) | 0.986 |
| **Cardiometabolic risk factors** |  |  |  |  |
| Body weight, kg | 89.8 (14.9) | 89.1 (14.8) | 91.8 (15.3) | 0.057 |
| Height, cm | 179.1 (8.1) | 178.9 (8.2) | 179.5 (7.9) | 0.494 |
| Waist circumference, cm | 104.1 (12.4) | 103.4 (12.6) | 106.1 (12.0) | 0.031 |
| Hip circumference, cm | 104.3 (8.8) | 104.0 (9.0) | 105.0 (8.8) | 0.242 |
| Systolic blood pressure, mm Hg | 131.2 (14.7) | 130.6 (14.6) | 132.6 (14.8) | 0.152 |
| Use of antihypertensive medication (%) | 234 (41.6) | 147 (36.5) | 87 (54.4) | < 0.001 |
| Use of diabetes medication (%) | 68 (12.1) | 35 (8.7) | 33 (20.6) | < 0.001 |
| Use of lipid-lowering medication (%) | 138 (24.5) | 81 (20.1) | 57 (35.6) | < 0.001 |
| Current smoker (%) | 27 (4.8) | 20 (5.0) | 7 (4.4) | 0.940 |
| Family history of myocardial infarction* (%) | 126 (22.4) | 84 (20.8) | 42 (26.2) | 0.202 |
| **Laboratory variables** |  |  |  |  |
| LDL cholesterol, mmol/L | 3.6 (1.1) | 3.7 (1.1) | 3.5 (1.2) | 0.014 |
| HDL cholesterol, mmol/L | 1.4 (0.3) | 1.4 (0.3) | 1.4 (0.3) | 0.846 |
| Triglycerides (mmol/L) | 1.2 [0.8, 1.7] | 1.2 [0.8, 1.7] | 1.1 [0.8, 1.6] | 0.383 |
| Total cholesterol, mmol/L | 5.3 (1.1) | 5.3 (1.1) | 5.4 (1.1) | 0.032 |
| Apolipoprotein B, g/L | 1.1 (0.3) | 1.0 (0.3) | 1.1 (0.3) | 0.039 |
| Lipoprotein(a) (g/L) | 0.11 [0.04, 0.34] | 0.11 [0.05, 0.32] | 0.11 [0.04, 0.36] | 0.838 |
| HbA1c, mmol/mol | 37.6 (7.4) | 37.1 (7.2) | 38.7 (7.7) | 0.024 |
| Plasma glucose, mmol/L | 6.0 (1.3) | 5.9 (1.3) | 6.1 (1.3) | 0.512 |
| GFR, mL/min/1.73 m<sup>2</sup> | 78.5 (13.5) | 78.3 (12.7) | 81.3 (15.2) | 0.448 |
| Creatinine, μmol/L | 80.5 (21.7) | 80.3 (19.4) | 81.3 (24.1) | 0.505 |
| High-sensitivity CRP (mg/L) | 1.3 [0.6, 2.70] | 1.3 [0.63, 2.60] | 1.4 [0.64, 3.30] | 0.434 |
| **Coronary calcium score** |  |  |  |  |
| CAC score ≥100 (%) | 28.4 | 0.0 | 100.0 |  |
| CAC score ≥300 (%) | 12.6 | 0.0 | 44.4 |  |

\* Father, mother, or biological sibling had a myocardial infarction before 60 years of age. Values are presented as n (%), mean (SD), or median [interquartile range (IQR)] as appropriate.

### Agreement between self-reported and clinical variables

Agreement between variables obtained from the initial web-based questionnaire and corresponding variables reassessed or measured during the clinical visit is presented in Supplemental Table S8. Predicted probabilities based on self-reported anthropometric variables were highly correlated with probabilities recalculated using clinically reassessed waist and hip circumference measurements among participants with available paired data (R=0.96, p<2.2e-16; Supplemental Figure S1).

### Agreement between predicted and observed CAC

Among participants who underwent CT imaging (n=563), the mean predicted probability of CAC ≥100 was 28.3% (95% CI 27.2–29.3), similar to the observed prevalence of 28.4% (95% CI 24.8–32.4) (Figure 2A). The expected/observed ratio was 0.99, with calibration intercept −0.11 and calibration slope 0.87. Calibration plots comparing predicted probability with observed prevalence of CAC ≥100 across tertiles and quintiles of predicted risk showed overall agreement across risk strata (Figures 2B–C). The Brier score was 0.19 and Spiegelhalter’s Z-test indicated no evidence of miscalibration (p=0.92).

**Figure 2:**
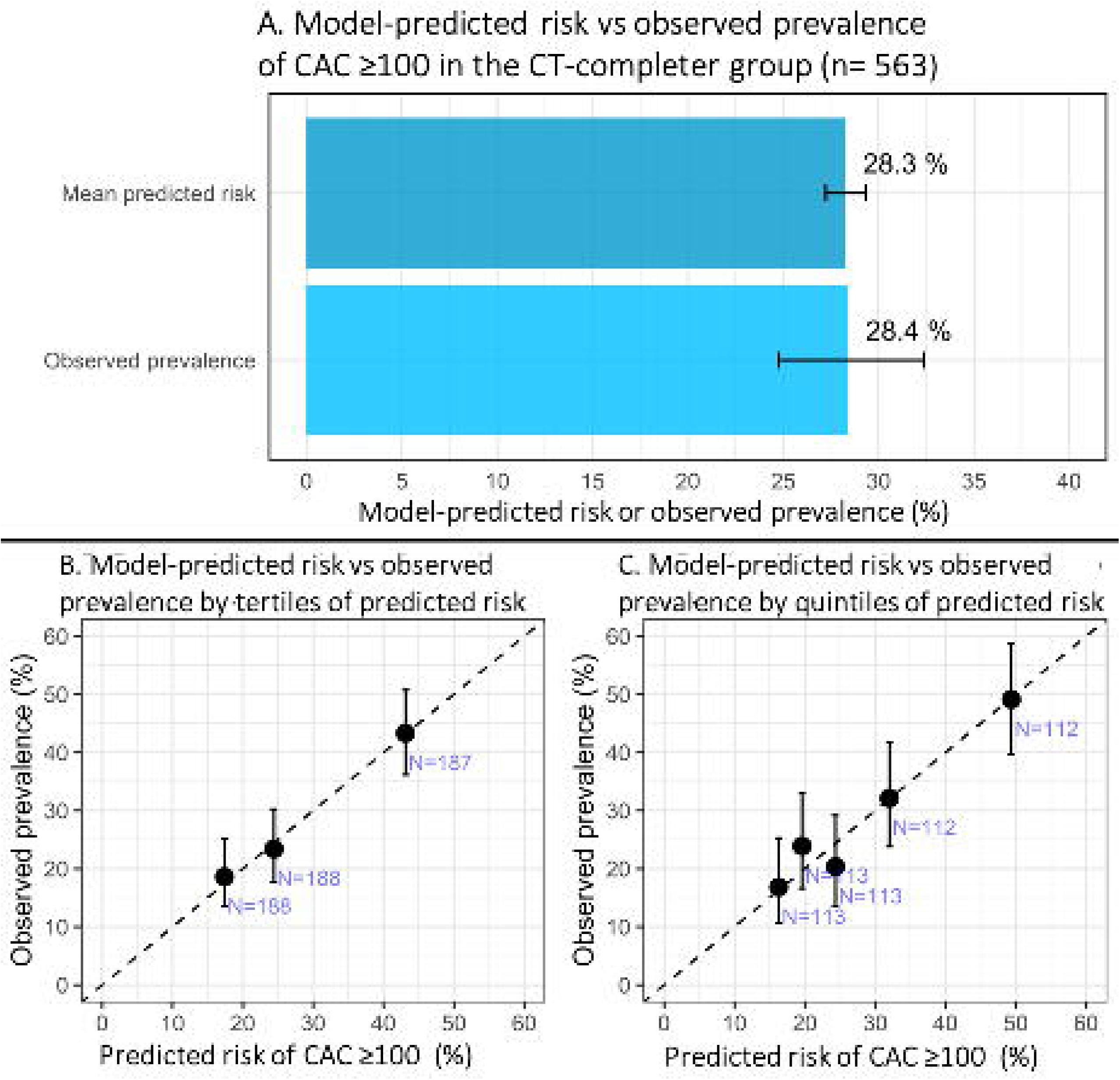
Model-predicted probability and observed prevalence of CAC ≥100 among participants who underwent CT imaging (n=563). **A.** Mean model-predicted probability and observed prevalence of CAC ≥100. Error bars represent 95% confidence intervals for mean model-predicted probability and observed prevalence. **B.** Calibration plot by tertiles of model-predicted probability. **C.** Calibration plot by quintiles of model-predicted probability. Error bars in panels B and C represent 95% confidence intervals for observed prevalence of CAC ≥100 within each risk group. CAC, coronary artery calcium; PRIO-Now, PRedict and Identify cOronary atherosclerosis-Now.

### Lipid-lowering treatment status among individuals with CAC ≥100 and ≥300

Descriptive distributions of lipid-lowering therapy use and LDL-C levels among individuals with CAC ≥100 and ≥300 are presented in Figures 3–4 and Tables 3–4. Among participants with CAC ≥100 (n=160), 64% were not receiving lipid-lowering therapy (Figure 3A). Among treated individuals, approximately 75% had LDL-C above 1.8 mmol/L, and only 8% were both treated and at the study-defined CAC-based target level (Figure 3B, Table 3). Among participants with CAC ≥300 (n=71), 61% were not receiving lipid-lowering therapy (Figure 4A). Only 4% had LDL-C levels ≤1.4 mmol/L, and 1.4% were both treated and at target (Figure 4B, Table 4).

**Figure 3:**
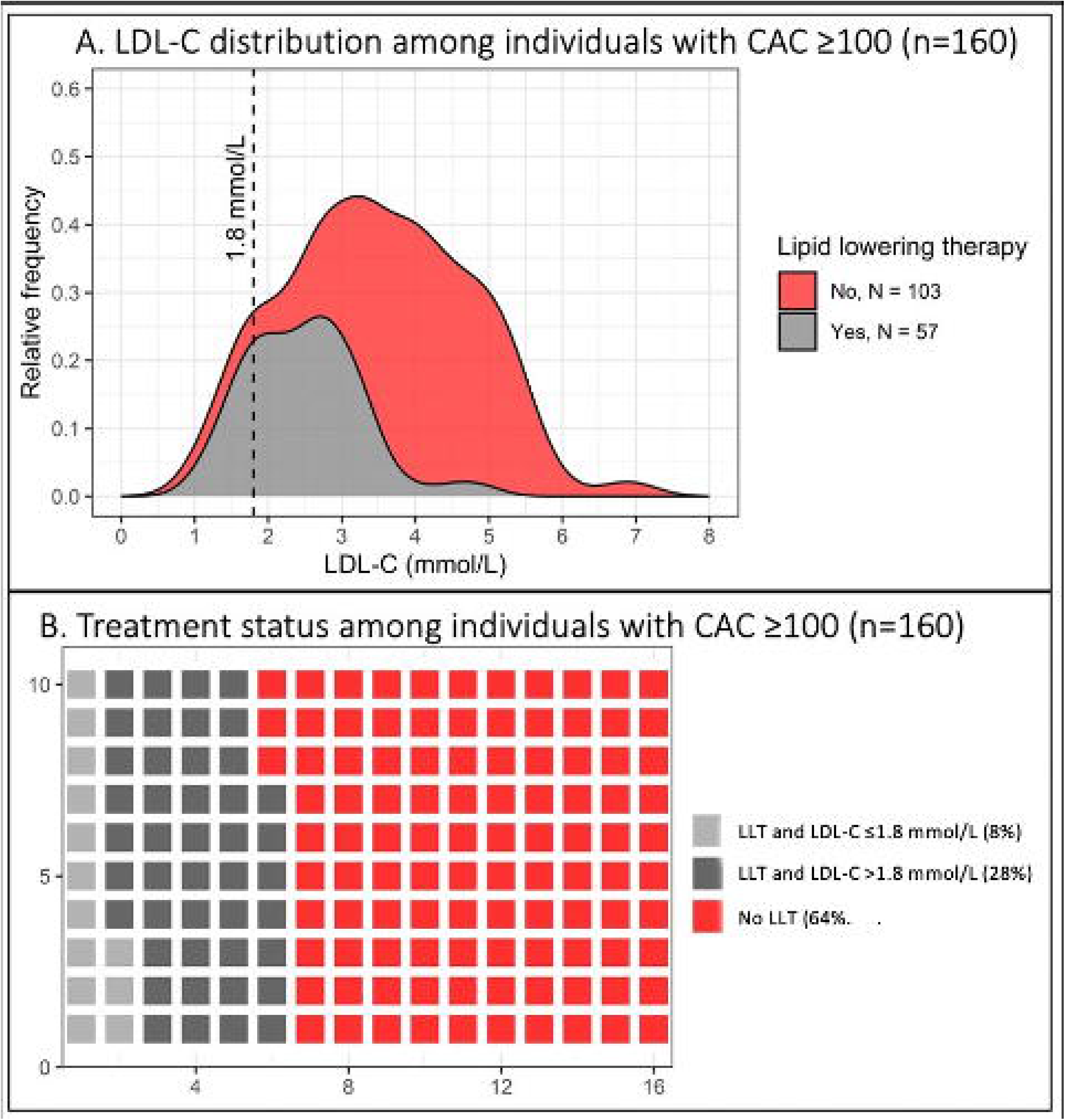
Treatment status among individuals with moderate to severe atherosclerosis (n=160, CAC ≥100). **A.** Distribution of LDL-C levels among individuals with CAC ≥100, stratified by lipid-lowering treatment status. **B.** Overview of lipid-lowering therapy status and LDL-C levels among participants with CAC ≥100. Each square represents one participant. Overall, 64% reported no lipid-lowering therapy, and 8% were receiving lipid-lowering therapy and had LDL-C ≤1.8 mmol/L. CAC: coronary artery calcium.

**Figure 4:**
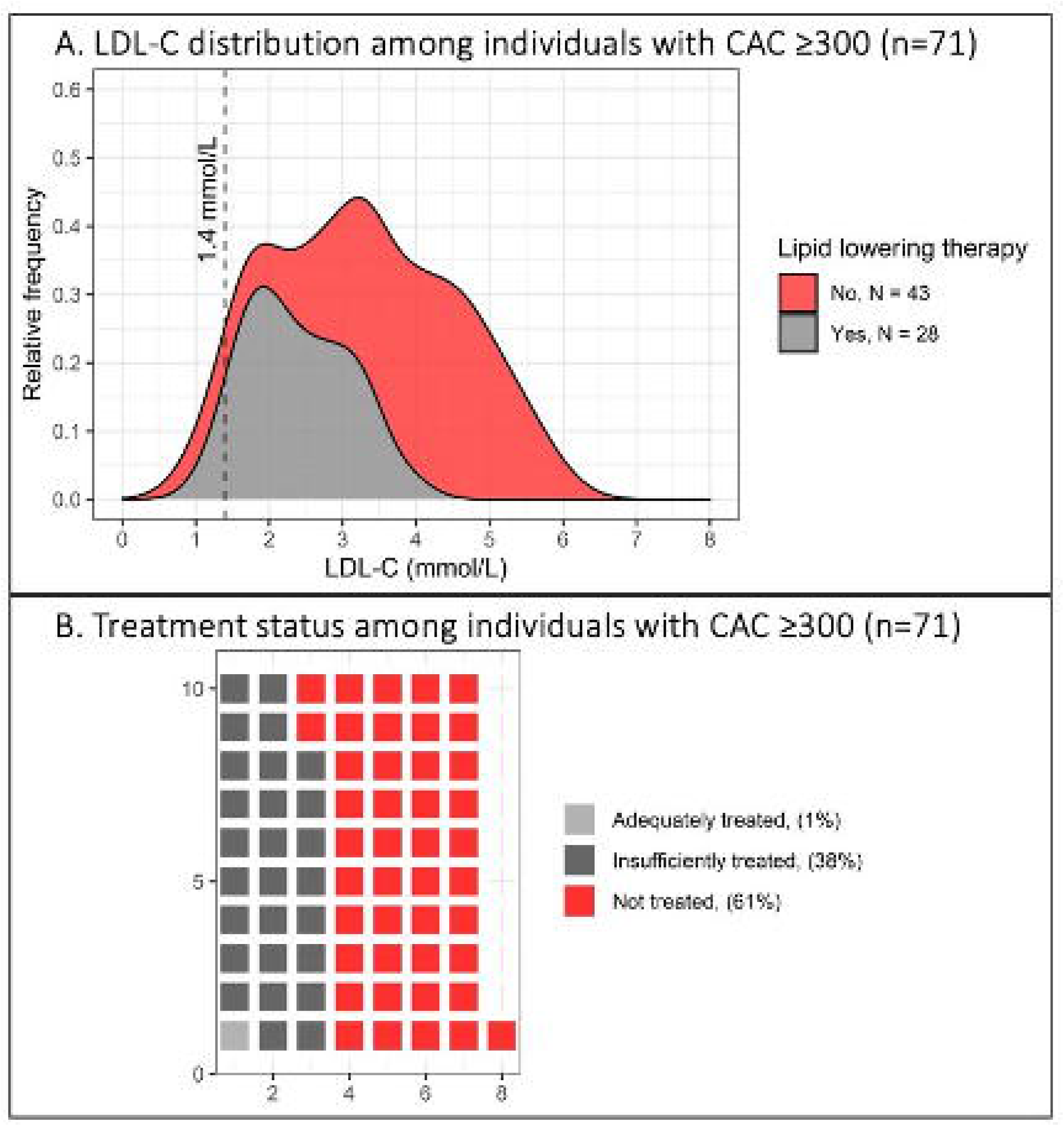
Treatment status among individuals with severe atherosclerosis (n=71, CAC ≥300). **A.** Distribution of LDL-C levels among individuals with CAC ≥300, stratified by lipid-lowering treatment status. B. Overview of lipid-lowering therapy status and LDL-C levels among participants with CAC ≥300. Each square represents one participant. Overall, 61% reported no lipid-lowering therapy, and 1.4% were receiving lipid-lowering therapy and had LDL-C ≤1.4 mmol/L. CAC: coronary artery calcium.

**Table 3:** LDL-C levels in untreated and treated participants with CAC ≥100 according to study-defined LDL-C levels.

| Characteristic | Total<br>(n=160) | Untreated<br>(n=103) | Treated<br>(n=57) |
| --- | --- | --- | --- |
| Mean LDL-C level, mmol/L | 3.5 (1.2) | 4.0 (1.1) | 2.5 (0.8) |
| LDL-C level $\leq 1.8$ mmol/L (n, %) | 18 (11.3%) | 5 (4.9%) | 13 (22.8%) |
| LDL-C level $> 1.8 - \leq 3.0$ mmol/L (n, %) | 39 (24.4%) | 9 (8.7%) | 30 (52.6%) |
| LDL-C level $> 3.0$ mmol/L (n, %) | 103 (64.4%) | 89 (86.4%) | 14 (24.6%) |

Values are presented as mean (SD) or n (%).

**Table 4:** LDL-C levels in untreated and treated participants with CAC ≥300 according to study-defined LDL-C levels.

| Characteristic | Total<br>(n=71) | Untreated<br>(n=43) | Treated<br>(n=28) |
| --- | --- | --- | --- |
| Mean LDL-C level, mmol/L | 3.3 (1.2) | 3.8 (1.2) | 2.4 (0.7) |
| LDL-C level $\leq 1.4$ mmol/L (n, %) | 3 (4.2%) | 2 (4.7%) | 1 (3.6%) |
| LDL-C level $> 1.4 - \leq 3.0$ mmol/L (n, %) | 28 (39.4%) | 7 (16.3%) | 21 (75.0%) |
| LDL-C level $> 3.0$ mmol/L (n, %) | 40 (56.3%) | 34 (79.1%) | 6 (21.4%) |

Values are presented as mean (SD) or n (%).

### Incidental findings

Incidental non-coronary radiological findings are summarised in Supplemental Table S9. Overall, 38.2% of participants had at least one incidental finding. Heart valve calcifications were observed in 22.0%, predominantly involving the aortic valve (101 cases). Ascending aortic dilation (>40 mm) was present in 14.5%, and hiatus hernias in 4.1%. Alterations in the lung parenchyma, including nodules and consolidations, were found in 7.5% of participants. Among participants with lung parenchymal abnormalities (n=42), 76% (n=32) were referred for follow-up thoracic CT imaging.

## Discussion

Non-contrast CT imaging enables detection of calcified coronary atherosclerosis, but its cost and resource implications favour targeted use in individuals with a sufficiently high pre-test probability.^7^ In this pragmatic prospective study, a two-step strategy using a self-report model to select individuals for CAC imaging showed close agreement between predicted risk and observed prevalence of CAC ≥100 among participants 59-60 year old completing CT imaging. Complementary calibration metrics supported this agreement. These findings suggest that self-report–based pre-selection may support targeted CAC imaging.

Most individuals with CAC ≥100 and ≥300 were not receiving lipid-lowering therapy, and LDL-C levels frequently remained above study-defined targets even among those treated. These findings are clinically relevant, as CAC is a robust marker of future cardiovascular risk, and higher CAC categories have consistently been associated with increased event rates in observational cohorts.^2-5,15,18^ Observational data further suggest that individuals with elevated CAC may experience greater relative risk reduction with lipid lowering therapy compared with those without calcification.^19^

CAC staging commonly uses thresholds of 1-99, 100-299, and ≥300 corresponding to mild, moderate, and severe calcified coronary artery atherosclerosis respectively.^14^ In the SCAPIS cohort, CAC ≥100 and CAC ≥300 were found in 11.6% and 4.7% of individuals aged 50–64 years, respectively.^7^ In both SCAPIS and other prospective cohorts, these CAC thresholds have been associated with a 3- to 6-fold increased hazard of cardiovascular events for CAC ≥100 and hazard ratios exceeding 10 for those with CAC ≥300.^18,20,21^ Given these markedly elevated risk levels, LDL-C targets of ≤1.8 mmol/L for individuals with CAC ≥100 and ≤1.4 mmol/L for CAC ≥300 have been proposed by specialty recommendations and recent expert consensus statements.^14,15,20^ Although not yet incorporated in major international guidelines,^15,16^ these targets reflect a shift toward more personalized lipid management in individuals with high CAC burden.

The PRIO-Now self-report–based questionnaire was designed to identify individuals most likely to benefit from CAC imaging. Previous analyses have demonstrated that the PRIO-Now model identifies approximately 64% of all individuals with CAC ≥100 in adults aged 50–64 years and performs favourably compared with guideline-based risk assessment.^9^ In a proof-of-concept analysis within the Multi-Ethnic Study of Atherosclerosis, the model provided a more resource-efficient and accurate approach to identifying individuals at risk of CHD compared with a strategy based on the Pooled Cohort Equation followed by CAC imaging.^22^

Self-report pre-screening has been explored in cardiovascular prevention research as a feasible population-based risk assessment strategy,^9,23,24^ but to our knowledge, this is the first prospective study evaluating self-report-based selection specifically for CAC imaging. The web-based format may facilitate large-scale use of self-reported pre-screening without the need for in-clinic risk assessment. In the present study, predicted probabilities based on self-reported waist and hip circumference were highly correlated with probabilities recalculated using clinically reassessed measurements, suggesting limited influence on model-based risk estimates.

The inclusion of participants receiving statin therapy may have influenced the observed CAC distribution, as statins are known to promote plaque calcification.^25^ However, the model was developed and calibrated in a population including statin users, and calibration metrics remained overall robust. LDL-C levels frequently exceeded study-defined targets even among treated individuals, suggesting potential room for treatment optimisation.

Non-contrast CT imaging for CAC also detects extracoronary findings. In this study, 5.7% of participants underwent follow-up thoracic CT imaging, consistent with previously reported re-scanning rates.^26^ Incidental findings should be considered in cost-effectiveness analyses of CAC screening strategies. Prior studies have suggested that CAC testing can be cost-effective^27^ and that CAC visualization may improve adherence to preventive therapies.^28^

The predominance of men in the model-selected high-risk group reflects the higher prevalence of coronary atherosclerosis at this age in men, whereas women typically develop coronary artery disease later.^7,29^ Future implementation may therefore require consideration of older age groups for women to achieve a more balanced identification of individuals with elevated CAC. The observed socioeconomic differences in response rates to the initial web-based questionnaire further highlight the importance of considering potential disparities in uptake when implementing questionnaire-based preventive screening strategies at the population level.

Future evaluation studies may further simplify the screening pathway. For example, clinical assessment and CT imaging could potentially be performed during the same visit to reduce participant burden and minimise attrition between screening steps. In addition, the use of mandatory questionnaire fields may reduce missing data.

### Limitations

This study has several limitations. The response rate was 32%, and the study population therefore represents a selected subset of invited individuals. Area-level socioeconomic and sex differences in response rates were observed, but other individual-level characteristics of non-responders were not available. Direct assessment of response bias was therefore limited. Non-response may have influenced both the distribution of predicted risk and the observed prevalence of CAC ≥100 among CT completers, and findings should be interpreted within the context of a model-selected study cohort rather than the full invited population.

Participant attrition also occurred after high-risk classification. Although baseline characteristics were largely similar between individuals who completed CT imaging and those who did not (Supplemental Table S5), some selection bias cannot be excluded.

Reliance on self-reported questionnaire data may introduce recall bias. Missing self-reported anthropometric variables may also have introduced some uncertainty in individual-level risk estimates, although the overall proportion of missing data was modest.

Finally, the high-risk group selected for CT imaging was predominantly male, likely reflecting the higher prevalence of CAC ≥100 among men in this age range. In addition, the narrow age range of 59–60 years, selected to reduce age-related variation in CAC prevalence, may limit generalisability to other age groups and broader population settings.

## Conclusion

In this prospective evaluation study, a self-report-guided strategy enabled targeted CAC imaging by selecting individuals at increased predicted risk of CAC ≥100. Among participants completing CT imaging, the observed prevalence of CAC ≥100 was comparable with the mean model-predicted probability. These findings suggest that simple self-report data may support pragmatic pre-selection for CAC imaging and help identify opportunities for preventive treatment optimisation among individuals with elevated CAC.

## Acknowledgments

The authors thank all study participants and the staff involved in the PRIO-Now study, as well as the Department of Radiology at Sahlgrenska University Hospital.

## Supplemental data

Supplemental Methods, Supplemental Tables S1–S9.

