## Supplementary Material for "Prospective evaluation of a self-report–guided strategy for targeted coronary artery calcium imaging"

**Supplemental Material**

| **Section** | **Page** |
| --- | --- |
| Supplemental Methods |  |
| Model development and application in PRIO-Now | 2 |
| Visit 1 – Clinical assessment | 2 |
| Visit 2 – Coronary CT imaging | 3 |
| Communication and referral process | 3 |
| Supplemental Table S1 – Overview of model inputs and measurement scales in the self-report model, with the Pooled Cohort Equation (PCE) and SCORE2 shown for comparison | 5 |
| Supplemental Table S2 – Missing data in self-reported variables | 6 |
| Supplemental Table S3 – Missing data in clinical variables | 6 |
| Supplemental Table S4 – Comparison of self-reported variables between participants who attended versus did not attend the clinical assessment **(step 2)** | 7 |
| Supplemental Table S5 – Comparison of baseline characteristics of participants who completed versus did not complete CT imaging | 8 |
| Supplemental Table S6 – Glucose control stratified by diabetes status | 8 |
| Supplemental Table S7 – Hypertension distribution stratified by treatment status | 9 |
| Supplemental Table S8 – Comparison between initial self-reported and clinically reassessed cardiovascular risk factors among participants attending the clinical visit (n=643) | 10 |
| Supplemental Table S9 – Secondary findings from coronary CT imaging | 11 |
| Supplemental Figure S1 – Predicted probabilities using self-reported versus clinically reassessed anthropometric variables | 12 |

**Model development and application in PRIO-Now
The prediction model was originally developed in the SCAPIS cohort** using extreme gradient boosting (XGBoost).^11^ The final model included 14 self-reported variables selected based on model performance and clinical relevance. The outcome was moderate to severe coronary atherosclerosis, defined as coronary artery calcium score (CAC) ≥100. Model development included 5-fold cross-validation, hyperparameter optimization and receiver operating characteristic curve analysis. Comparisons were performed using the DeLong test. External validation was performed in an independent SCAPIS subcohort (n=1,111).

For the present study, the previously published risk threshold corresponding to the 30% at highest-risk was applied.¹¹ In the SCAPIS population aged 59–60 years, this threshold corresponded to a predicted probability of 15.3%. The threshold and the model were not changed when moved to the Prio-Now population.

**Visit 1 – Clinical assessment**
All self-reported data from the pre-screening questionnaire were verified at the clinical visit. Anthropometric measurements, including waist circumference, were obtained and participants completed a detailed electronic case report form covering self-reported health status, medication use, lifestyle factors, psychosocial well-being, socioeconomic variables, and family history. Medication lists were reviewed and documented by a trained study nurse.

Blood pressure was measured three times using an automated oscillometric device (Omron M10-IT, Omron Health Care Co, Kyoto, Japan), with the average of the last two measurements used.

Fasting venous blood samples were collected for laboratory analyses. Glucose metabolism was assessed using plasma glucose (p-Glucose), glycated hemoglobin (HbA1c) and capillary glucose (Hemocue). Lipid profile analyses included triglycerides (s-TG), total cholesterol (s-Cholesterol), LDL-C, HDL-C, apolipoprotein B (ApoB), and lipoprotein(a). Inflammatory status was evaluated using high-sensitivity C-reactive protein (hs-CRP). Standard haematological and liver function parameters were also measured according to established protocols.

**Visit 2 – Coronary CT imaging**
CAC imaging was performed using a dual-source Somatom Force CT scanner (Siemens Medical Solutions, Germany). CAC scores were calculated using syngo.via CT CaScoring software, following manual verification of coronary segmentation by a CT-trained radiology nurse.

**Communication and referral process**
Participants with predicted risk ≥15.3% received a personalised written report summarising results from clinical assessment and CT imaging, including laboratory values, anthropometric measures, blood pressure, and CAC score where available. Reports included reference ranges and explanatory text to support interpretation, and participants were advised that findings were non-diagnostic and should be discussed with their healthcare provider.

Blood pressure levels <140/90 mm Hg were recommended, with 120/80 mm Hg described as an optimal level for individuals at elevated cardiovascular risk. Participants with HbA1c ≥42 mmol/mol were advised to seek clinical follow-up.

Participant feedback included study-defined LDL-C levels according to CAC burden and additional risk factors, with suggested levels of <3.0 mmol/L for all participants, <1.8 mmol/L for CAC ≥100, and <1.4 mmol/L for CAC ≥300. Participants with diabetes were advised to discuss LDL-C levels with their primary care provider. Individuals with CAC ≥300 were additionally contacted by telephone, and treatment recommendations were discussed with a cardiologist.

All CT scans were reviewed by a radiologist, and incidental extracoronary findings were documented and evaluated by the study physician. Participants were informed of clinically relevant findings and advised regarding follow-up where appropriate. Participants who did not undergo CT imaging received feedback based on clinical assessment alone, including cardiovascular risk evaluation and lifestyle recommendations.

**Supplemental Table S1**

Overview of model inputs and measurement scales in the PRIO-Now model, with SCORE2 and the Pooled Cohort Equation (PCE) shown for comparison.

| **Category** | **Factor** | **PRIO-Now (scale)** | **SCORE2 (scale)** | **PCE**  **(scale)** |
| --- | --- | --- | --- | --- |
| Demographics | Sex | Binary | Binary | Binary |
|  | Age  Ethnicity | Continuous  N/A | Continuous  N/A | Continuous  Categorical (Black/White) |
| Anthropometry | Body weight at age 20 | Continuous | N/A | N/A |
|  | Body weight | Continuous | N/A | N/A |
|  | Height | Continuous | N/A | N/A |
|  | Waist circumference | Continuous | N/A | N/A |
|  | Hip circumference | Continuous | N/A | N/A |
| Smoking status | Smoking | Binary | Binary | Binary |
|  | Cigarette pack-years | Continuous | N/A | N/A |
| Health condition | Use of lipid-lowering medication | Binary | N/A | N/A |
|  | Use of antihypertensive medication | Binary | Binary | Binary |
|  | Diagnosed diabetes | Binary | N/A | Binary |
|  | Diagnosed hypertension | Binary | N/A | N/A |
| Blood tests | Total cholesterol | N/A | Continuous | Continuous |
|  | HDL cholesterol | N/A | N/A | Continuous |
| Clinical assessment | Systolic blood pressure | N/A | Continuous | Continuous |
| Family history | Family history of myocardial infarction* | Binary | N/A | N/A |

N/A, Not Applicable. * Father, mother, or biological sibling had a myocardial infarction before 60 years of age.

**Supplemental Table S2**

Missing data in self-reported variables

| **Variable** | **Missing** | **Total** | **Missing, %** |
| --- | --- | --- | --- |
| Sex | 0 | 2375 | 0 |
| Age | 0 | 2375 | 0 |
| Body weight at age 20 | 98 | 2375 | 4.1 |
| Current body weight | 100 | 2375 | 4.2 |
| Height | 131 | 2375 | 5.5 |
| Waist circumference | 385 | 2375 | 16.2 |
| Hip circumference | 415 | 2375 | 17.5 |
| Current smoker | 0 | 2375 | 0 |
| Pack-years of smoking | 7 | 2375 | 0.3 |
| Lipid-lowering medication use | 0 | 2375 | 0 |
| Diagnosed diabetes | 91 | 2375 | 3.8 |
| Diagnosed hypertension | 91 | 2375 | 3.8 |
| Family history of myocardial infarction* | 92 | 2375 | 3.9 |

* Father, mother, or biological sibling had a myocardial infarction before 60 years of age.

**Supplemental Table S3**

Missing data in clinical variables

| **Variable** | **Missing** | **Total** | **Missing %** |
| --- | --- | --- | --- |
| Apolipoprotein B (g/L) | 4 | 643 | 0.6 |
| Triglycerides (mmol/L) | 2 | 643 | 0.3 |
| Lipoprotein(a) (g/L) | 2 | 643 | 0.3 |
| LDL cholesterol (mmol/L) | 0 | 643 | 0.0 |
| HDL cholesterol (mmol/L) | 0 | 643 | 0.0 |
| Total cholesterol (mmol/L) | 0 | 643 | 0.0 |
| hs-CRP (mg/L) | 10 | 643 | 1.6 |
| Plasma glucose (mmol/L) | 1 | 643 | 0.2 |
| HbA1c (mmol/mol) | 0 | 643 | 0.0 |
| Creatinine (µmol/L) | 1 | 643 | 0.2 |
| GFR (mL/min/1.73 m²) | 2 | 643 | 0.3 |

**Supplemental Table S4**

Comparison of self-reported variables between participants who attended versus did not attend the clinical assessment **(step 2)**

| Characteristics | Overall  (n=814) | Attended step 2 (n=643) | Did not attend step 2 (n=171) | p-value |
| --- | --- | --- | --- | --- |
| Sex (n, %, male) | 709 (87.1) | 570 (88.6) | 139 (81.3) | 0.015 |
| Age, years | 59.5 (0.58) | 59.5 (0.6) | 59.4 (0.6) | 0.708 |
| Body weight at 20, kg | 74.0 (10.9) | 74.3 (10.6) | 73.1 (12.1) | 0.199 |
| Body weight, kg | 89.4 (15.4) | 89.8 (15.0) | 88.2 (17.0) | 0.226 |
| Height, cm | 178.6 (8.3) | 179.0 (8.2) | 177.0 (8.8) | 0.008 |
| Waist circumference, cm | 103.9 (12.8) | 104.2 (12.4) | 102.6 (14.3) | 0.208 |
| Hip circumference, cm | 104.1 (10.0) | 104.4 (9.1) | 102.8 (13.3) | 0.109 |
| Current smoker (%) | 68 (8.4) | 45 (7.0) | 23 (13.5) | 0.011 |
| Cigarette pack-years, median [IQR] | 0.00  [0.00, 13.00] | 0.0 [0.0, 12.0] | 1.7  [0.0, 18.5] | 0.021 |
| Use of lipid-lowering medication (%) | 191 (23.5) | 150 (23.3) | 41 (24.0) | 0.939 |
| Use of antihypertensive medication (%) | 337 (41.4) | 266 (41.4) | 71 (41.5) | 1.000 |
| Diagnosed diabetes (%) | 92 (11.3) | 74 (11.5) | 18 (10.5) | 0.822 |
| Diagnosed hypertension (%) | 449 (55.2) | 359 (55.8) | 90 (52.6) | 0.508 |
| Family history of myocardial infarction (%)* | 161 (19.8) | 135 (21.0) | 33 (19.3) | 0.626 |

* Father, mother, or biological sibling had a myocardial infarction before 60 years of age.

Values are presented as n (%), mean (SD), or median [interquartile range (IQR)] as appropriate.

**Supplemental Table S5**

**Comparison of baseline characteristics of participants who** completed versus did not complete CT imaging **(n=643)**

| Variable | Overall,  n= 643 | With CT,  n= 563 | Without CT,  n=80 | p-value |
| --- | --- | --- | --- | --- |
| Sex (n, % male) | 570 (88.6) | 500 (88.8) | 70 (87.5) | 0.875 |
| Age (years ± SD) | 59.5 (0.6) | 59.5 (0.6) | 59.5 (0.6) | 0.926 |
| Body weight (kg ± SD) | 89.8 (15.0) | 89.8 (15.0) | 89.3 (15.1) | 0.762 |
| Height (cm ± SD) | 179.0 (8.2) | 179.1 (8.1) | 178.1 (8.4) | 0.294 |
| Waist circumference (cm ± SD) | 104.3 (12.6) | 104.1 (12.4) | 105.1 (13.9) | 0.535 |
| Hip circumference (cm ± SD) | 104.4 (9.1) | 104.3 (8.8) | 105.6 (11.0) | 0.271 |
| LDL cholesterol, mmol/L | 3.7 (1.1) | 3.7 (1.1) | 3.5 (1.0) | 0.251 |
| HDL cholesterol, mmol/L | 1.4 (0.3) | 1.4 (0.3) | 1.3 (0.3) | 0.253 |
| Triglycerides (mmol/L), median [IQR] | 1.20  [0.84, 1.70] | 1.20  [0.83, 1.70] | 1.30  [0.90, 1.70] | 0.365 |
| HbA1c (mmol/mol ± SD) | 37.5 (7.1) | 37.6 (7.4) | 36.8 (5.1) | 0.360 |
| Plasma glucose, mmol/L | 6.0 (1.3) | 6.0 (1.2) | 5.9 (1.3) | 0.500 |
| Total cholesterol, mmol/L | 5.3 (1.2) | 5.3 (1.2) | 5.1 (1.1) | 0.195 |
| Apolipoprotein B, g/L | 1.0 (0.3) | 1.0 (0.3) | 1.0 (0.2) | 0.248 |
| Lipoprotein(a) (g/L), median [IQR] | 0.11  [0.05, 0.36] | 0.11  [0.04, 0.34] | 0.12  [0.06, 0.46] | 0.179 |
| GFR, mL/min/1.73 m² | 78.5 (10.1) | 78.5 (10.3) | 78.0 (8.7) | 0.655 |
| Creatinine, µmol/L | 80.5 (16.6) | 80.5 (17.0) | 80.63 (13.2) | 0.956 |
| High-sensitivity CRP (mg/L), median [IQR] | 1.30  [0.67, 2.80] | 1.30  [0.63, 2.70] | 1.50  [0.83, 2.80] | 0.134 |
| Use of diabetes medication (%) | 75 (11.7) | 68 (12.1) | 7 (8.8) | 0.406 |
| Use of lipid-lowering medication (%) | 150 (23.3) | 138 (24.5) | 12 (15.0) | 0.521 |
| Use of antihypertensive medication (%) | 266 (41.4) | 234 (41.6) | 32 (40.0) | 0.663 |
| Cigarette pack-years, median [IQR] | 0.00  [0.00, 12.00] | 0.00  [0.00, 11.25] | 0.25  [0.00, 20.75] | 0.172 |

Values are presented as n (%), mean (SD), or median [interquartile range (IQR)] as appropriate.

**Supplemental Table S6**

Glucose control stratified by diabetes status among participants attending clinical assessment (n=643)

|  | **Total** | **<42 mmol/mol** | **42-47 mmol/mol** | **≥48 mmol/mol** |
| --- | --- | --- | --- | --- |
| No diagnosed diabetes, n (%) | 561 | 535 (95.4%) | 8 (1.4%) | 18 (3.2%) |
| With diabetes + Treated, n (%) | 75 | 16 (21.3%) | 38 (50.7%) | 21 (28.0%) |
| With diabetes + Untreated, n (%) | 7 | 5 (71.4%) | 1 (14.3%) | 1 (14.3%) |
| Total, n (%) | 643 | 556 (86.5%) | 47 (7.3%) | 40 (6.2%) |

**Supplemental Table S7**

Hypertension distribution by treatment status among participants attending clinical assessment (n=643)

|  | **Total** | **<140 mm Hg (%)** | **≥140 mm Hg (%)** |
| --- | --- | --- | --- |
| No diagnosed hypertension, n (%) | 284 | 232 (81.7%) | 52 (18.3%) |
| With hypertension + Treated, n (%) | 266 | 183 (68.8%) | 83 (31.2%) |
| With hypertension + Untreated, n (%) | 93 | 59 (63.4%) | 34 (36.6%) |
| Total | 643 | 474 (73.7%) | 169 (26.3%) |

**Supplemental Table S8**

Comparison between initial self-reported and clinically reassessed cardiovascular risk factors among participants attending the clinical visit (n=643)

| Variable | Self-reported questionnaire | Clinical visit | Difference | p-value |
| --- | --- | --- | --- | --- |
| Body weight, kg | 89.8 (15.0) | 90.2 (15.3) | +0.4 | <0.001 |
| Waist circumference, cm | 104.2 (12.4) | 101.3 (12.0) | −2.9 | <0.001 |
| Hip circumference, cm | 104.4 (9.1) | 104.0 (8.7) | −0.4 | 0.005 |
| Body weight at age 20 (kg) | 74.3 (10.6) | 74.2 (11.2) | −0.1 | 0.641 |
| Current smoking (%) | 45 (7.0) | 55 (8.6) | +1.6 pp | 0.016 |
| Diagnosed hypertension (%) | 359 (55.8) | 354 (55.1) | −0.7 pp | 0.532 |
| Use of antihypertensive medication (%) | 266 (41.4) | 270 (42.0) | +0.6 pp | 0.386 |
| Use of lipid-lowering medication (%) | 150 (23.3) | 151 (23.5) | +0.2 pp | 1.000 |
| Diagnosed diabetes (%) | 74 (11.5) | 81 (12.6) | +1.1 pp | 0.023 |
| Family history of myocardial infarction | 133 (20.7) | 117 (18.2) | −2.5 pp | 0.034 |

Values are presented as mean (SD) or n (%), as appropriate. Differences represent clinical visit minus self-reported values; for categorical variables, differences are shown as percentage-point (pp) differences. Analyses were based on available paired data.

**Supplemental Table S9**

Secondary incidental findings from coronary CT imaging

| Secondary findings | Total, n | Percentage of total scans % | Percentage of all incidental findings % |
| --- | --- | --- | --- |
| Ascending aorta dilatation | 82 | 14.6 | 27.2 |
| *40-45 mm* | *74* | 13.1 | 24.5 |
| *>45 mm* | *8* | 1.4 | 2.6 |
| Descending aorta dilatation | 1 | 0.2 | 0.3 |
| Pulmonary trunk dilatation | 1 | 0.2 | 0.3 |
| Other heart anomalies/alterations | 2 | 0.4 | 0.7 |
| Heart valve calcifications | 124 | 22.0 | 41.1 |
| Aortic valve | 101 | 17.9 | 33.4 |
| Mitral valve | 23 | 4.1 | 7.6 |
| Mediastinal lymph node enlargement/calcification | 10 | 1.8 | 3.3 |
| Esophagus alteration | 4 | 0.7 | 1.3 |
| Lung parenchyma | 42 | 7.5 | 13.9 |
| Nodule | 18 | 3.2 | 6.0 |
| Condensation | 12 | 2.1 | 4.0 |
| Other alterations in the lung parenchyma | 12 | 2.1 | 4.0 |
| Hiatus hernia | 25 | 4.4 | 8.3 |
| Liver – cysts/calcifications | 8 | 1.4 | 2.6 |
| Bone – hypoattenuating lesion | 1 | 0.2 | 0.3 |
| Cutaneous/subcutaneous lesions | 2 | 0.4 | 0.7 |
| Total secondary findings | 302 |  |  |
| Participants with incidental findings | 215 | 38.2 |  |
| Participants without incidental findings | 348 | 61.8 |  |
| Total | 563 | 100 |  |

**Supplemental Figure S1**
Predicted probabilities using self-reported versus clinically reassessed anthropometric variables

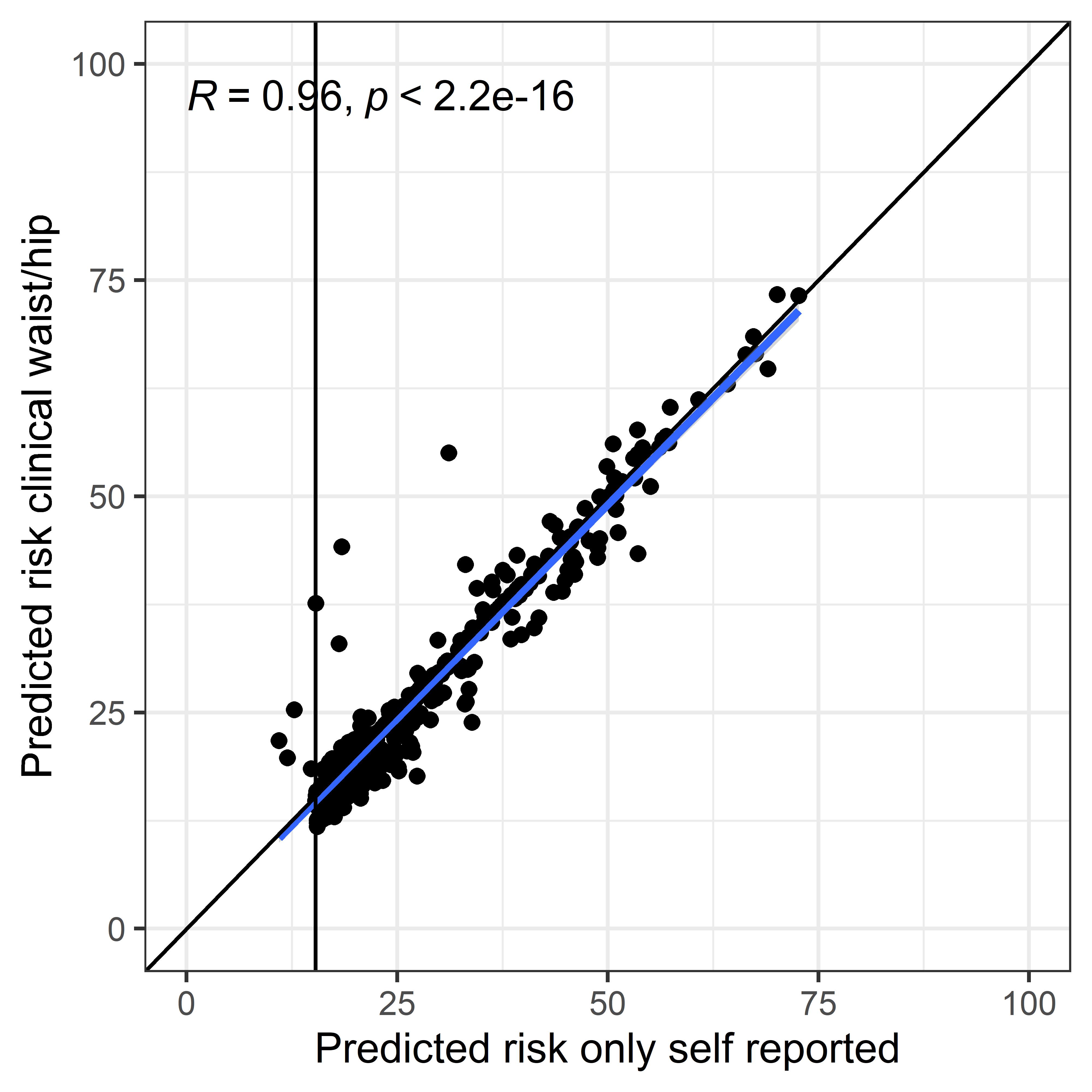

Correlation between predicted probabilities derived using self-reported waist and hip circumferences and probabilities recalculated using clinically reassessed measurements among participants with available paired data. The vertical line indicates the predefined high-risk threshold of 15.3%; the diagonal line represents the line of identity.
